# *“This is about the coolest thing I’ve ever seen is that you just showed right up.”* COVID-19 testing and vaccine acceptability among homeless-experienced adults: Qualitative data from two samples

**DOI:** 10.1101/2021.03.16.21253743

**Authors:** Kelly Ray Knight, Michael R. Duke, Caitlin A. Carey, Graham Pruss, Cheyenne M. Garcia, Marguerita Lightfoot, Elizabeth Imbert, Margot Kushel

## Abstract

**Background:** Homeless-experienced populations are at increased risk of exposure to SARS CoV-2 due to their living environments and face increased risk of severe COVID-19 disease due to underlying health conditions. Little is known about COVID-19 testing and vaccination acceptability among homeless-experienced populations.

**Objective:** To understand the facilitators and barriers to COVID-19 testing and vaccine acceptability among homeless-experienced adults.

**Design:** We conducted in-depth interviews with participants from July-October 2020. We purposively recruited participants from 1) a longitudinal cohort of homeless-experienced older adults in Oakland, CA (n=37) and 2) a convenience sample of people (n=57) during a mobile outreach COVID-19 testing event in San Francisco.

**Participants:** Adults with current or past experience of homelessness.

**Approach:** We asked participants about their experiences with and attitudes towards COVID-19 testing and their perceptions of COVID-19 vaccinations. We used participant observation techniques to document the interactions between testing teams and those approached for testing. We audio-recorded, transcribed and content analyzed all interviews and identified major themes and subthemes.

**Key Results:** Participants found incentivized COVID-19 testing administered in unsheltered settings and supported by community health outreach workers (CHOWs) to be acceptable. The majority of participants expressed positive inclination toward vaccine acceptability, citing a desire to return to routine life and civic responsibility. Those who expressed hesitancy cited a desire to see trial data, concerns that vaccines included infectious materials, and mistrust of the government.

**Conclusions:** Participants expressed positive evaluations of the incentivized, mobile COVID-19 testing supported by CHOWs in unsheltered settings. The majority of participants expressed positive inclination toward vaccination. Vaccine hesitancy concerns must be addressed when designing vaccine delivery strategies that overcome access challenges. Based on the successful implementation of COVID-19 testing, we recommend mobile delivery of vaccines using trusted CHOWs to address concerns and facilitate wider access to and uptake of the COVID vaccine.

## Introduction

Approximately 550,000 people are homeless each night in the United States; Black and Native Americans are overrepresented. [1] Adults experiencing homelessness, particularly those living in congregate shelters, are at high risk of acquiring SARS-CoV-2 infection due to environmental conditions (i.e. crowded spaces, poor ventilation). [2] Behavioral conditions common in homeless populations (i.e. mental health disorders) can interfere with preventive behaviors (e.g. mask wearing, social distancing) that reduce the risk of SARS-CoV-2 transmission. There have been multiple large outbreaks of COVID-19 in homeless shelters. [2]

Homeless-experienced adults have a high prevalence of conditions associated with severe COVID-19 illness, including chronic lung and kidney disease, and cancer. [3] They have low rates of insurance, low use of primary care, and high rates of Emergency Department use and hospitalization. [4, 5] They face barriers to early intervention if they are infected. For these reasons, some states have prioritized people experiencing homelessness for COVID-19 vaccination. [6]

Many factors (i.e. inability to travel far distances, lack of primary care physician, transient movement) contribute to homeless populations being difficult to reach for COVID-19 testing and vaccination, and reportedly could decrease acceptability of these interventions. [5] Due to experiences of stigma and racism in health settings, homeless populations may also mistrust health care interventions. [7, 8] Limited internet access [9] may decrease their ability to receive information about or register to receiver testing and vaccines. Little is known about homeless-experienced adults’ attitudes towards and willingness to accept COVID-19 testing or vaccination. We conducted in-depth interviews with currently and formerly homeless individuals (homeless-experienced) to understand their experience with and attitudes towards testing and vaccination to inform strategies to improve the delivery and uptake of COVID-19 testing and vaccination in this population.

## Methods

### Setting and Study Population

We recruited from two separate homeless-experienced samples. For one, we recruited 37 participants from the Health Outcomes in People Experiencing Homelessness in Older Middle agE (“HOPE HOME”) study, a longitudinal study of people ≥50 years old who were experiencing homelessness at enrollment (July 2013 to June 2014 and August 2017 to June 2018) in Oakland, CA. [10, 11] We used purposive sampling to recruit HOPE HOME participants from three current living categories: congregate shelters or unsheltered settings (n=12), pandemic-response non-congregate shelters (“shelter-in-place” (SIP) hotels) (n=12), or housed (n=13). [10]

For the second sample, we recruited 57 participants during a two-day COVID-19 mobile testing event in October 2020 for unsheltered individuals in San Francisco, CA. We used convenience sampling to recruit individuals who did (n=50) and did not (n=7) test. Before the event, Community Health Outreach Workers (CHOWs) advertised the event to unsheltered individuals. CHOW-led testing teams spread out throughout the geographic areas with the highest number of homeless-experienced adults. [12] The teams offered testing in homeless encampments, on street corners, and in front of a large homeless services organization. All who tested received an incentive of a cloth mask, snacks, and $10 gift card.

### Data Collection and Analysis

Interviewers conducted hour-long HOPE HOME qualitative interviews by telephone in July and August 2020. Interview topics included participants’ experiences of and health-related behaviors during the COVID-19 pandemic, access to health care, and acceptability of influenza and COVID-19 vaccines.

During the testing event, ethnographers accompanied testing teams and recruited individuals for 20-minute interviews. The interview examined perceptions of, and prior experiences with, COVID-19 testing; impressions of mobile versus stationary COVID-19 testing; opinions about isolation and quarantine options provided by San Francisco to people infected with COVID-19; and acceptability of influenza and COVID-19 vaccines. Prior to the testing event, ethnographers documented the CHOWs’ outreach efforts. During the event, ethnographers documented their observations of interactions between the testing team and participants.

We provided participants with gift cards ($25 for HOPE HOME and $20 for testing event). All interviews were audio-recorded and professionally transcribed. We developed a codebook for the HOPE HOME interviews through an iterative, consensus process, and coded interview transcripts via Dedoose. We repeated this process for the shorter mobile testing interviews with a condensed codebook. We conducted content analysis on interviews and fieldnote observations. The University of California, San Francisco’s Institutional Review Board approved all study procedures for both projects.

## Results

Of the 94 participants, two-thirds (65%) were cis-men and 3% were transgender. Over half (56%) identified as Black, 24% as white, 7% as Latinx, and 4% as Native American, Asian/Pacific Islander, and other, respectively. Participants ranged in age from 20-71 with a median age of 59. (Table 1)

**Table 1.**
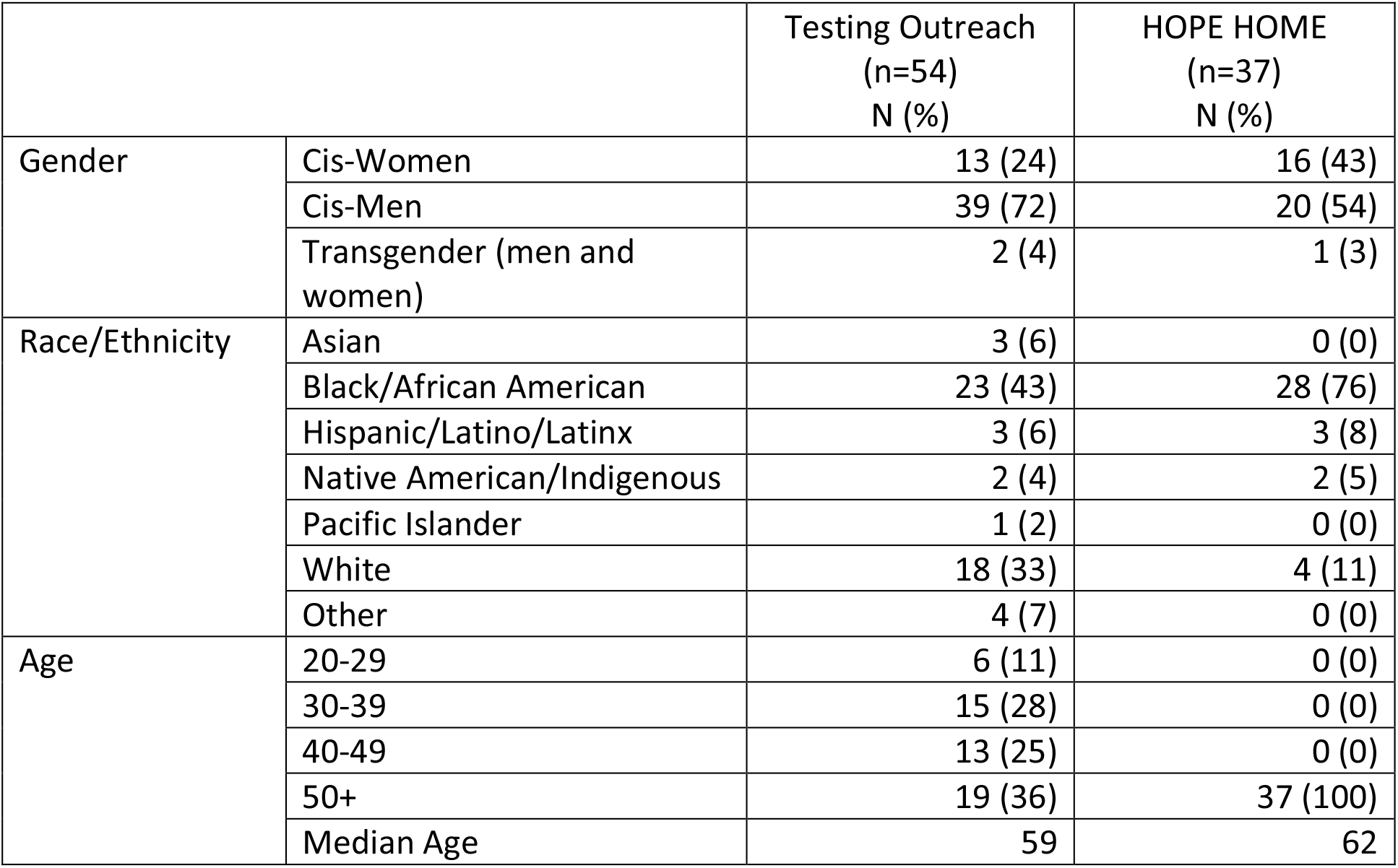
Sample Characteristics

| Table 1. Sample Characteristics |  | Testing Outreach<br>(n=54)<br>N (%) | HOPE HOME<br>(n=37)<br>N (%) |
| --- | --- | --- | --- |
| Gender | Cis-Women | 13 (24) | 16 (43) |
|  | Cis-Men | 39 (72) | 20 (54) |
|  | Transgender (men and women) | 2 (4) | 1 (3) |
| Race/Ethnicity | Asian | 3 (6) | 0 (0) |
|  | Black/African American | 23 (43) | 28 (76) |
|  | Hispanic/Latino/Latinx | 3 (6) | 3 (8) |
|  | Native American/Indigenous | 2 (4) | 2 (5) |
|  | Pacific Islander | 1 (2) | 0 (0) |
|  | White | 18 (33) | 4 (11) |
|  | Other | 4 (7) | 0 (0) |
| Age | 20-29 | 6 (11) | 0 (0) |
|  | 30-39 | 15 (28) | 0 (0) |
|  | 40-49 | 13 (25) | 0 (0) |
|  | 50+ | 19 (36) | 37 (100) |
|  | Median Age | 59 | 62 |

Facilitators for testing were the convenience of mobile testing, support for incentives, recognition of COVID-19’s severity, and support for universal testing. Barriers to testing were fears of shelter disruption and concerns about the accuracy and safety of testing. (Table 2) For COVID-19 vaccine acceptability, we found the desire to reunite with family and work, and a sense of civic responsibility to be motivating factors, and a desire to see safety and efficacy data, prior negative experience with vaccines, desire to wait for others to be vaccinated first, and mistrust of the government as reasons for vaccine hesitancy. (Table 3)

**Table 2.** Themes for COVID-19 Testing and Mobile Outreach

|  |
| --- |
| **Testing Facilitators** |
| **Theme 1. Convenience of Mobile Testing** |
| *"[I learned about the COVID testing event] cause you walked up to me." [Testing event participant]* |
| *"That's [mobile testing] the only way, I mean, like I move around so much." [Testing event participant]* |
| **Theme 2. Incentives** |
| *"I mean, serious, like I don't want to catch it [COVID-19]... I don't want to catch anything but some money." [Testing event participant]* |
| *"Probably announce giftcards. They [other people experiencing homelessness] won't do it [COVID-19 testing] if it's like... for free." [Testing event participant]* |
| **Theme 3. Concerns about COVID-19 severity and symptoms** |
| *"I just don't want to get sick, period. I'm a person that don't need to get sick, with having the problems that I already have I don't need to get sick." [HOPE HOME participant]* |
| *"[What prevented you from getting tested?] I just, I just feel normal, I don't know." [HOPE HOME participant]* |
| **Theme 4. Support for Universal Testing** |
| *"Well I think that, uh, people that work in restaurants and fast food restaurants should be tested even if they say they had it cause they should send someone down there from the health department and test them anyway. Because you never know. They don't want to miss a paycheck and so they're probably not even getting tested, you know." [Testing event participant]* |
| *"I know it sounds horrible and kind of like not very privacy oriented, but I still think mandatory testing would be a good idea." [Testing event participant]* |
| **Testing Barriers** |
| **Theme 5. Fear of Shelter Disruption** |
| *"It [moving into an isolation and quarantine hotel] really upset me, and this is the second time that I've had all my things... I had to give up so many, I had bought clothes, buy food, everything, I spent close to \$500 on getting supplies when I first hear about the epidemic and I had all new thing, I hadn't even had it two days and they gave me the test and put me in the hotel. And the hotel said I had to leave all my stuff..." [HOPE HOME participant]* |
| **Theme 6. Concerns about Accuracy and Safety** |
| *"The scary part about that is half the people in that line (for testing) are already sick. So you're telling me you want me to stand in line with somebody who's sick, and no guarantee that they're going to stay their distance from me? ... I see the testing sites but I'm afraid to go stand in it because I may stand in it and actually get it." [HOPE HOME Participant]* |

**Table 3.**
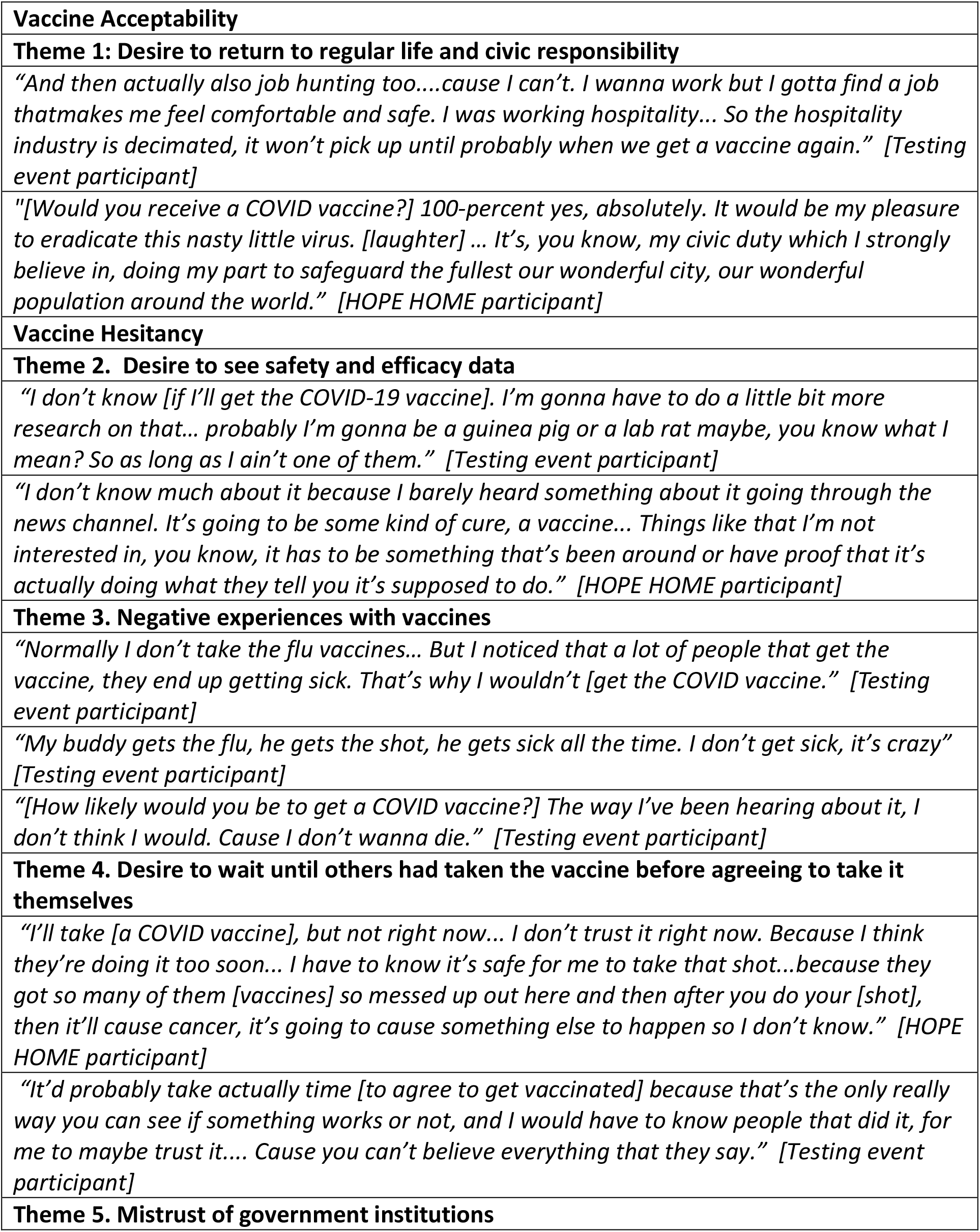
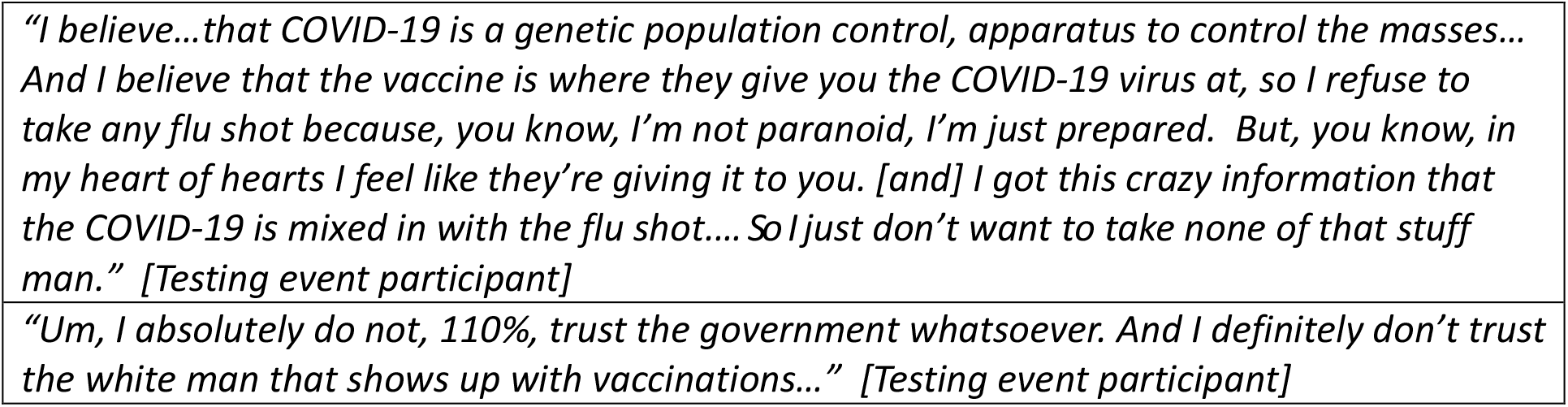
Themes for COVID-19 Vaccines

|  |
| --- |
| **Vaccine Acceptability** |
| **Theme 1: Desire to return to regular life and civic responsibility** |
| *"And then actually also job hunting too....cause I can't. I wanna work but I gotta find a job that makes me feel comfortable and safe. I was working hospitality... So the hospitality industry is decimated, it won't pick up until probably when we get a vaccine again." [Testing event participant]* |
| *"[Would you receive a COVID vaccine?] 100-percent yes, absolutely. It would be my pleasure to eradicate this nasty little virus. [laughter] ... It's, you know, my civic duty which I strongly believe in, doing my part to safeguard the fullest our wonderful city, our wonderful population around the world." [HOPE HOME participant]* |
| **Vaccine Hesitancy** |
| **Theme 2. Desire to see safety and efficacy data** |
| *"I don't know [if I'll get the COVID-19 vaccine]. I'm gonna have to do a little bit more research on that... probably I'm gonna be a guinea pig or a lab rat maybe, you know what I mean? So as long as I ain't one of them." [Testing event participant]* |
| *"I don't know much about it because I barely heard something about it going through the news channel. It's going to be some kind of cure, a vaccine... Things like that I'm not interested in, you know, it has to be something that's been around or have proof that it's actually doing what they tell you it's supposed to do." [HOPE HOME participant]* |
| **Theme 3. Negative experiences with vaccines** |
| *"Normally I don't take the flu vaccines... But I noticed that a lot of people that get the vaccine, they end up getting sick. That's why I wouldn't [get the COVID vaccine]." [Testing event participant]* |
| *"My buddy gets the flu, he gets the shot, he gets sick all the time. I don't get sick, it's crazy" [Testing event participant]* |
| *"[How likely would you be to get a COVID vaccine?] The way I've been hearing about it, I don't think I would. Cause I don't wanna die." [Testing event participant]* |
| **Theme 4. Desire to wait until others had taken the vaccine before agreeing to take it themselves** |
| *"I'll take [a COVID vaccine], but not right now... I don't trust it right now. Because I think they're doing it too soon... I have to know it's safe for me to take that shot...because they got so many of them [vaccines] so messed up out here and then after you do your [shot], then it'll cause cancer, it's going to cause something else to happen so I don't know." [HOPE HOME participant]* |
| *"It'd probably take actually time [to agree to get vaccinated] because that's the only really way you can see if something works or not, and I would have to know people that did it, for me to maybe trust it.... Cause you can't believe everything that they say." [Testing event participant]* |
| **Theme 5. Mistrust of government institutions** |

*"I believe...that COVID-19 is a genetic population control, apparatus to control the masses... And I believe that the vaccine is where they give you the COVID-19 virus at, so I refuse to take any flu shot because, you know, I'm not paranoid, I'm just prepared. But, you know, in my heart of hearts I feel like they're giving it to you. [and] I got this crazy information that the COVID-19 is mixed in with the flu shot.... So I just don't want to take none of that stuff man." [Testing event participant]*
*"Um, I absolutely do not, 110%, trust the government whatsoever. And I definitely don't trust the white man that shows up with vaccinations..." [Testing event participant]*

### Testing Facilitators

#### Theme 1: Convenience of Mobile Testing

Many participants noted that mobile testing was convenient and acceptable.

> *“This is about the coolest thing I’ve ever seen is that you just showed right up. On-the-spot testing*…*on the sidewalk like that, it was just a good idea*.*” [Testing event participant]*

Mobile teams were convenient because they reached people who did not want to abandon their belongings or leave their neighbors to participate. Study ethnographers observed community members thanking the teams for bringing incentivized testing to where they lived. Ethnographers documented that CHOWs initiated conversations and built rapport with participants before and during the event. CHOWs’ familiarity with participants aided in managing expectations and resolving tensions between participants waiting to test.

#### Theme 2: Incentives

In this low-income population, participants emphasized the importance of the $10 gift card and food as facilitators to testing. Almost every participant noted the importance of incentives.

> *“You can’t make it any easier, because you’re paying them [to test] now, so I’d probably keep on testing as much as I can*.*” [Testing event participant]*

#### Theme 3: Concerns about COVID-19 severity and symptoms

Participants noted that they were concerned about the severity of COVID-19 and its interaction with their underlying health conditions. This concern motivated some to seek out testing.

> *“I actually am high risk too because I have co-morbidities. So, if I catch [COVID] I’m at risk for severe illness*…*Diabetes, high blood pressure and cholesterol. The trifecta’*.*” [Testing event participant]*

Some participants noted that they would test if they had COVID-like symptoms. Conversely, participants who declined to test cited their lack of symptoms as a reason.

> *“If I felt kind of sick I would have gone for the test*…*but I don’t feel sick or anything*…*I feel okay*.*” [HOPE HOME participant]*

#### Theme 4: Support for universal testing

Some participants voiced support for broader testing requirements, including universal testing.

> *“Whether you have the symptoms or not, I think everybody should be tested*… *I don’t care if you have to get an appointment or whatever, everybody needs to be tested*.*” [HOPE HOME participant]*

### Testing Barriers

#### Theme 5: Fear of shelter disruption

Participants noted a testing barrier was fear of losing access to their spots in sanctioned encampments or shelters if they tested positive.

> *“If you disappear, then there’s a good reason as to why you’re disappearing. ‘He left all his stuff?, Oh, he must have COVID”*…*Even the staff would be like, “He had that [COVID], I don’t know if we want to let him back in here’*…*I feel like, even if I went into isolation and didn’t have [COVID anymore], that would happen. So, just to be on the safe side, it’s like the backlash from doing the right thing. It’s gonna have a serious effect on me*.*” [Testing event participant]*

#### Theme 6: Concerns about accuracy and safety

Participants expressed concerns that the COVID-19 test could give someone the virus, by exposing them to others while testing and by direct inoculation during testing.

> *“[People] think they’re getting tested and they could be given the virus*…*Because they have ways of giving people this virus and they don’t know how they’re getting it, but that’s one way they can do that*.*” [HOPE HOME participant]*
>
> Some were concerned that the tests could produce false results that would cause harm.

> *“Some of the testing that they’ve had, they’ve had false positives and I’m just like, ‘No, I’m not going to do that*…*I’m not sure about the reliability of the tests, to be honest with you*.*” [HOPE HOME participant]*

### Vaccine Acceptability

#### Theme 1: Desire to return to routine life and civic responsibility

Despite the interviews taking place prior to the release of trial data showing vaccine efficacy and vaccines receiving Emergency Use Authorization (EUA), many participants indicated a willingness to be vaccinated. They cited a desire to reunite with family, work and engage in everyday activities as reasons to be vaccinated.

> *“[I would] definitely [get the COVID vaccine]. ‘Cause I can’t be around my kids right now. My mom’s not letting me come around my baby girl. [S]he has strict like rules on me*…*if I don’t meet them*…*she will definitely not let me go in the house*.*” [Testing event participant]*

Others were motivated by a sense of civic responsibility.

> *“We got to get this epidemic handled*…*If they come up with a vaccine that actually works, I think everybody should get it because we got to get rid of this*…*Because you can’t have some people getting a vaccine and then you’re missing [other] people*…*Everybody got to have it if we want to get rid of this*… *I think that’s the biggest challenge right there, how is you going to get everyone vaccines?” [HOPE HOME participant]*

### Vaccine Hesitancy

#### Theme 2: Desire for data

Participants who expressed an inclination to take the vaccine nevertheless noted the need for more data about vaccine testing, safety, and approval.

> *“I would probably take that vaccine*…*I would be very aware and conscious of the safety of it*…*If it’s been proved by the USDA [sic] for it being a safe vaccine, yes, I would*.*” [Testing event participant]*

#### Theme 3: Negative experiences with other vaccines

Participants expressed concerns that the vaccines made people sick. This stemmed from observing reactions to other vaccines and believing that vaccine side effects (e.g. sore arms, low grade fevers) or other seasonal illnesses were signs that vaccines led to illness. Some expressed beliefs that vaccines contained a live virus that led to these symptoms.

> *“It seems like every time you take a vaccine, you get sick. And so my understanding is vaccines are made from the viruses themselves. I don’t want you to* …*inject the virus into me, to keep me from getting the virus. And if I don’t have it now, I’m going to leave it alone*.*” [HOPE HOME participant]*

#### Theme 4: Desire to wait until others had taken the vaccine before agreeing to take it themselves

Some wanted to see public figures or trusted community members vaccinated first, expressing concerns about being the first to receive a vaccine.

> ***“****I wouldn’t want to take (the vaccine) unless I saw somebody like Donald Trump and some of the other people taking it, until they did it and the statistics came back. I wouldn’t want to be the guinea pig guy. Nope*.*” [HOPE HOME participant]*

#### Theme 5: Mistrust of government institutions

Participants who expressed concerns about vaccines noted their mistrust of government.

> *“I truly don’t trust the government a lot. There’s just a lot of weird stuff going on, it seems*.*” [Testing event participant]*

Some connected this mistrust to experiences of racism.

> *“I’m a Black person and I’m kind of skeptical about [being among the first to get the vaccine], because, as ethical as they sound and as transparent as they’re supposed to be and all of this stuff, I know that CDC and the medical department, they’ve been known to experiment on people. “[HOPE HOME participant]*

## Discussion

In this qualitative study of two samples of homeless-experienced adults, we found that although homeless-experienced individuals face numerous barriers to accessing health services, offering low-barrier mobile testing mediated by CHOWs and incentivized by a small monetary gift and food led to testing acceptability and uptake. Despite conducting the interviews prior to the release of vaccine trial efficacy data and subsequent EUA by the Food and Drug Administration [13], we found a general willingness to receive the vaccine. This was motivated by an awareness of COVID-19’s severity, the desire to return to regular life, and a sense of civic responsibility. Many participants expressed the need to see safety data and trusted community members’ acceptance of the vaccine prior to agreeing to be vaccinated. Those who expressed hesitancy noted several concerns, similar to those about testing: the possibility that the vaccine could cause illness, skepticism about safety of the vaccine, and mistrust of the government.

Participants recognized the importance of testing but faced barriers to receive it. For example, there is a no-cost walk-in testing site in the neighborhood where we did our mobile testing. Despite this, participants still expressed the importance of the mobile outreach, which didn’t require them to leave belongings unattended or make appointments. People experiencing homelessness face numerous barriers to COVID-19 information, testing, and vaccination, including transportation impediments and lack of engagement in routine healthcare. [4, 5] Thus, the use of mobile COVID testing teams was a welcomed testing facilitator for individuals because it can mitigate these barriers. [14] Participants noted incentives were an important factor in their decision to test.

Participants noted that incentives counteract the cost of lost time due to testing when they have competing demands for basic survival. Incentives are an accepted strategy to nudge people who are hesitant about vaccinations. [15] We found support for the use of homeless-experienced CHOWs with neighborhood familiarity to facilitate outreach and increase testing uptake. Trained by, and working in partnership with, healthcare providers, the CHOWs brought evidence-based recommendations to the population and relayed questions to the healthcare team. This model offers an effective strategy for vaccinating this population. The CHOWs can extend the health care providers’ role by serving as trusted health messengers. They can identify concerns that need to be addressed, reduce operational barriers by guiding mobile outreach, and share their own narratives of vaccination to increase acceptability.

Concerns about testing mirror those about vaccinations and provide lessons. Some expressed concern that testing could transmit SARS-CoV-2; others had the same concern about vaccination. These concerns may be due to misinformation or prompted by a lack of trust in the healthcare system. Trusted messengers, such as CHOWs, should provide information to counteract these concerns. Others expressed the concern that testing could incur negative consequences. Vaccine campaigns should be cautious about unintended negative impacts, such as losing access to COVID19-response long-term non-congregate shelter. Policies should disconnect vaccinations from decisions to scale back these interventions, to reduce the chance that individuals sense that vaccine acceptance is tied to negative consequences.

Those who expressed support for vaccines noted their desire to return to regular life and a sense of civic responsibility. These reasons are in line with the general public and could inform the development of messaging for people experiencing homelessness. Participants also expressed an interest in examining safety data prior to vaccination. Despite their social exclusion and lack of access to technology, participants followed news reports about the vaccine and sought information about vaccine efficacy and safety. Efforts to increase vaccine uptake in this population should include easy to understand data about efficacy and safety.

Those who were hesitant about vaccines were concerned that the vaccine could give them COVID-19. They cited side effects of other vaccines as evidence that vaccines contained the disease. This apprehension echoed what some had expressed about testing. This concern may be amplified by observing people experiencing fevers, fatigue and other symptoms that may mimic COVID-19 after receiving the vaccine. [16] Trusted messengers should share information on the composition of the vaccine, anticipate concerns about expected side effects, reassuring homeless individuals that these symptoms are not a sign of the virus.

Some participants expressed hesitancy because they feared being experimental subjects and wanted to see public figures and peers vaccinated first. Some expressed mistrust of the government. These concerns underscore degrees of medical mistrust among people experiencing homelessness in the United States stemming from historic and ongoing racist medical policies and institutional structures. However, hesitancy does not equate to unwillingness. [17] Addressing medical racism directly through acknowledgement of its history and ongoing impact and providing examples of trusted public figures and community members getting vaccinated, could address some hesitancy. [18]

Our study has several limitations. We conducted our interviews prior to the release of vaccine study data and the resultant EUA. [13] We were not able to assess whether these events changed views. Additionally, in the testing sample, we were only able to interview a small number of those who chose not to be tested, so we may have understated objections to testing among the target population. Lastly, we reported data from two different studies whose populations were distinct in terms of age, testing accessibility and housing situation. This could also be a strength, since the similarity of responses between the two samples increases generalizability.

In a qualitative study of homeless-experienced individuals, we found that participants were interested in COVID-19 testing and vaccines and found CHOW-mediated mobile outreach to be effective and acceptable. Participants’ concerns about the vaccine mirrored those of the US general public. [19] However, homeless individuals face structural barriers stemming from a lack of housing coupled with a disproportionate burden of structural racism and discrimination based on social status and behavioral health characteristics. Our data suggest that providing mobile testing, incentives, and trusted members of communities (CHOWs) to provide information and answer questions can mitigate barriers to access and uptake of both COVID testing and vaccines.

## Data Availability

Not applicable due to the nature of our qualitative interviews. Data may contain identifying information.

## Acknowledgements

The authors would like to thank the participants for telling their stories. The authors would like to thank the CHOWS who worked with us and our community partners for their generous support during our testing events: Code Tenderloin, Glide Foundation, Larkin Street Youth Services, St. Anthony’s Foundation, and Urban Alchemy, and our partners for the HOPE HOME Study: St. Mary’s Center, Allen Temple, and Lifelong Medical Care. We could not do this work without the leadership and assistance of our Community Advisory Board. We would like to thank the hundreds of volunteers who helped us complete the mobile testing. Finally, the authors would also like to thank Deborah Yip and Ali Zahir for interviewing the HOPE HOME participants and Ashley Smith for coding the interviews.

## Funding Sources

This work was supported by the National Institute on Aging at National Institute of Health under Grants 5R01AG050630, 5R01AG041860, 2K24AG046372 awarded to Dr. Kushel, the Benioff Homelessness and Housing Initiative, and grants from Heluna Health and East Bay Community Foundation for the testing events.

The NIH had no role in the data collection, analysis or writing of the manuscript. The contents and views in this manuscript are those of the authors and should not be construed to represent the views of the National Institutes of Health.

